# The efficacy of Tripterygium Glycosides in the treatment of Chinese patients with thyroid-associated orbitopathy: A systematic review and meta-analysis

**DOI:** 10.1101/2024.06.18.24309103

**Authors:** Mingzhe Li, Bingchen Wei, Tianshu Gao, Chenghan Gao

## Abstract

**Objective:** This study aims to conduct a systematic review of the effectiveness and safety of Tripterygium Glycosides interventions in the treatment of Chinese patients with thyroid-associated orbitopathy (TAO).

**Methods:** A literature search was conducted using PubMed for English sources, and the CNKI, Chinese Biomedical Database, Wanfang Database, and VIP Database for Chinese sources. The search period extended from the beginning of the databases’ creation to Dec. 2023. The keywords used in the search were hyperthyroidism, thyroid-related immune orbitopathy (TRIO), ophthalmopathy, and Tripterygium Glycosides. Various combinations of search terms were used, depending on the database being queried. All the trials included in the study were clinical randomized controlled trials (RCTs).

**Results:** 33 RCTs or quasi-RCTs that met the inclusion criteria were included. The meta-analysis included 27 RCTs. 6 RCTs were excluded from the analysis due to the absence of a control group, but they were still included in the systematic review. 27 RCTs or quasi-RCTs involving 2,134 patients were included in the meta-analysis. The TRIO patients in the treatment group received Tripterygium Glycosides in combination with Thiamazole, Prednisone, Levothyroxine sodium, or a combination of these medications. While the TRIO patients in the control group were treated with Thiamazole, Prednisone, Levothyroxine sodium, or a combination of these treatments, the meta-analysis results show that the overall effectiveness rate of the treatment group and the control group was *P* = 0.05, I^2^ = 0.33 < 0.5 [MD = 4.45, 95% CI (3.31, 5.99), *P* < 0. 00001]. The former was significantly superior to the latter. At the same time, a risk assessment was conducted for the study of the 2 groups. The former was significantly superior to the latter. Furthermore, the clinical effectiveness rate of eyeball prominence was *P* < 0. 00001, I^2^ = 0.98 > 0.5 [MD = 2.40, 95% CI (2.28, 2.51), *P* < 0. 00001]. The clinical effectiveness rate of CAS score was *P* < 0. 00001, I^2^ = 0.89 > 0.5 [MD = 1.68, 95% CI (1.50, 1.85), *P* < 0. 00001]. The clinical effectiveness rate of FT_3_ was *P* < 0. 00001, I^2^ = 0.98 > 0.5 [MD = 0.95, 95% CI (0.81, 1.08), *P* < 0. 00001], the clinical effectiveness rate of FT_4_ was *P* < 0. 00001, I^2^ = 0.95 > 0.5 [MD = 2.12, 95% CI (1.99, 2.25), *P* < 0. 00001], and the clinical effectiveness rate of TSH was *P* < 0. 00001, I^2^ = 0.89 > 0.5 [MD = −0.19, 95% CI (−0.21, −0.17), *P* < 0. 00001].

**Conclusion:** The experience with the treatment of TAO using Tripterygium Glycosides was promising. The existing evidence suggests that treatment with Tripterygium Glycosides may be more effective in enhancing the response rate, quality of life, and FT_3_ levels compared to treatment with Prednisone, Levothyroxine sodium, and/or Thiamazole alone.

## 1. Introduction

Thyroid-associated ophthalmopathy (TAO) is a group of autoimmune diseases involving orbital and periocular tissues associated with genetic, environmental, and immunologic factors, with the highest incidence of orbital disease. The pathogenesis of the disease is complex, with the majority of patients suffering from Graves’ disease (GD), which has a prevalence of up to 40%. Moreover, in 80% of patients experiencing both hyperthyroidism and ophthalmopathy, the clinical symptoms progress rapidly within 2 years of disease onset, forming a vicious cycle [1]. Graves’ ophthalmopathy is also known as thyroid eye disease (TED), thyroid-associated orbitopathy (TAO), and Graves’ orbitopathy (GO) [2,3]. Tripterygium wilfordii is the Chinese herbalanti-inflammatory immunomodulator, which is the first studied andused in China, known as the “Chinese herbal hormone”. It has thefunctions of promoting blood circulation and collateralization, dispelling wind and dehumidification, detumescence and pain,detoxification, anti-inflammatory and etc. Extract of tripterygium wilfordii is often used in the treatment of autoimmune diseases. There is a large number of clinical studies having found thattripterygium wilfordii can be used in the treatment ofthyroid-associated ophthalmopathy [4]. Currently, the use of Tripterygium and its extracts for treating hyperthyroidism exophthalmos is gaining clinical attention. Comprehensive analysis and evaluation of RCTs on TAO with Tripterygium Glycosides were carried out in this paper according to principles of evidence-based medicine. A meta-analysis was conducted to provide objective and accurate evidence, and to assess the effectiveness of Tripterygium Glycosides in treating hyperthyroidism exophthalmos. The aim was to offer guidance and a foundation for the clinical use of this medication.

Tripterygium wilfordii is a perennial vine species in the Celastraceae family, extensively utilized in traditional Chinese medicine for the treatment of autoimmune and inflammatory diseases. According to the *Compendium of Materia Medica*, Tripterygium wilfordii is documented as a treatment for conditions such as swelling, edema, accumulation, yellow and white pox, long-term incurable malaria, constipation, leprosy, and falls.

The protocol of this network meta-analysis was registered in PROSPERO with ID CRD42021247873. We present the following article in accordance with the PRISMA reporting checklist (available at https://dx.doi.org/10.21037/apm-21-1307).

## 2. Methods

### 2.1. Literature sources and search

The publications utilized in the meta-analysis were identified through searches of the China National Knowledge Infrastructure (CNKI), PubMed, Web of Science, Wanfang Database, VIP Database, and EMBASE. The search period extended from the inception of the databases’ construction to December 2023, and the search was conducted in Chinese or English. The key words used in the search were “Hyperthyroidism”, “Thyroid related immune orbitopathy” or “TRIO”, “Ophthalmopathy” or “Tripterygium Glycosides”, “Tripterygium, Tripterygium wilfordii”, “Tripterygium wilfordii Hook f.”, “Tripterygium wilfordii multiglycoside”. Different combinations of search terms were used, depending on the selected database. The selected publications were clinical trials published in medical journals. 2 reviewers independently evaluated English and Chinese literature for inclusion. Any disagreements were resolved through discussion.

#### 2.1.1. Literature selection

Research on the types of RCTs (RCT or Controlled Clinical Trial, CCT) for the treatment of TAO, regardless of whether blinded or not.

Diagnose standard according to the *Diagnosis of clinical diseases based on the improvement of the standard* [5]: (1) typical ocular symptoms; (2) with hyperthyroidism or a history of hyperthyroidism; and (3) excluding other similar diseases.

Exclusion criteria: (1) myopia; (2) orbital inflammatory pseudotumor; (3) carotid-cavernous sinus fistula or dural artery cavernous sinus; (4) extraocular muscle lymphatic tumor; (5) primary orbital tumor; (6) ocular metastasis; and (7) intracranial tumors and other diseases.

TAO classification according to Wilmar’s simple classification standard. Grade Ⅰ (mild): eyeball prominence < 18mm, with upper eyelid retraction, gaze, eyelid, and conjunctival edema; Grade Ⅱ (moderate): eyeball prominence is 18-20 mm, with ocular involvement; Grade Ⅲ (severe): eyeball prominence > 20 mm, with corneal involvement and vision disorders [6].

#### 2.1.2. Literature extraction

Studies were excluded if they were: (1) animal experiments; (2) clinical trials from which no relevant data could be extracted; (3) repeatedly published studies; (4) studies involving patients with serious mental disorders or dementia; (5) studies involving patients with serious systemic symptoms that may significantly affect their ability to perform daily living activities, including syncope or coma, seizure-like headache, and cachexia; and (6) studies involving pregnant or breastfeeding women.

All trials included in the analysis were extracted by two reviewers. Once completed, any disagreements regarding data extraction and study evaluation were resolved through discussion with the third reviewer. All the trials included in the analysis contain information on study design, patient characteristics, and medication use.

### 2.2. Clinical efficacy

Clinical efficacy judgement [7,8]: Cure is defined as the disappearance of eye symptoms, obvious retraction of the eye, protrusion of the eyeballs < 18 mm, or a reduction in protrusion by > 3 mm. The treatment was significantly effective as the eye symptoms disappeared, but the reduction in exophthalmos > 2 mm. The degree of reduction in exophthalmos is effective, ranging from 1-2 mm. Invalid: The degree of exophthalmos showed no obvious change, or exophthalmos reduced by < 1 mm.

### 2.3. Quality assessment

Following the quality assessment standard recommended by the Cochrane Review Handbook 5.0 [9]. The bias risk assessment tool involved six aspects: (1) random distribution method; (2) concealment of allocation decisions; (3) blinding of research subjects, operators of the therapeutic plan, or those measuring the results; (4) result integrity; (5) selective presentation of study findings; and (6) other potential sources of bias.

Each research result was evaluated based on the six aspects mentioned above and categorized as “YES” (low-degree bias), “NO” (high-degree bias), or “unclear” (lacking relevant information or uncertain bias condition). Two evaluators cross-verified the quality assessment results of the inclusive trials, and any differences in opinions were resolved through discussion or third-party arbitration.

### 2.4. Statistical analysis

Meta-analysis was performed using the Rev Man software (Version 5.3) from The Cochrane Collaboration website. First, we performed clinical heterogeneity and methodological heterogeneity analyses for all the trials included. Statistical heterogeneity was evaluated by the Chi-squared (χ^2^) test and heterogeneity were considered present if *P* ≤ 0.10. A quantitative assessment of heterogeneity was performed using I^2^ where I^2^ > 50% indicated high heterogeneity among study results. Study results were pooled for analysis using a fixed effects model when there was no statistical heterogeneity or using a random effects model when statistical heterogeneity was detected. For dichotomous variables, odds ratio (OR) and 95% confidence interval (CI) was determined. For hypothesis testing, the U test was used and the results were presented as *Z* and *P* values. The differences in the efficacy between interventions were considered statistically significant if *P* ≤ 0.05. The results of hypothesis testing are presented in a forest plot.

## 3. Results

### 3.1. Identified studies and characteristics

The literature search yielded a total of 211 published studies. The abstracts of these studies were reviewed, and subsequently, 142 studies were excluded due to a lack of controls. The 69 potentially relevant RCTs were further reviewed, of which 36 were excluded due to the low Jaded score. Finally, 33 RCTs or quasi-RCTs that met the inclusion criteria were included. 27 RCTs were included in the meta-analysis, while 6 RCTs were excluded due to the absence of a control group, but they were included in the systematic review (Fig 1). A total of 33 RCTs with a diagnosis of TAO were included (Table 1).

**Fig 1.**
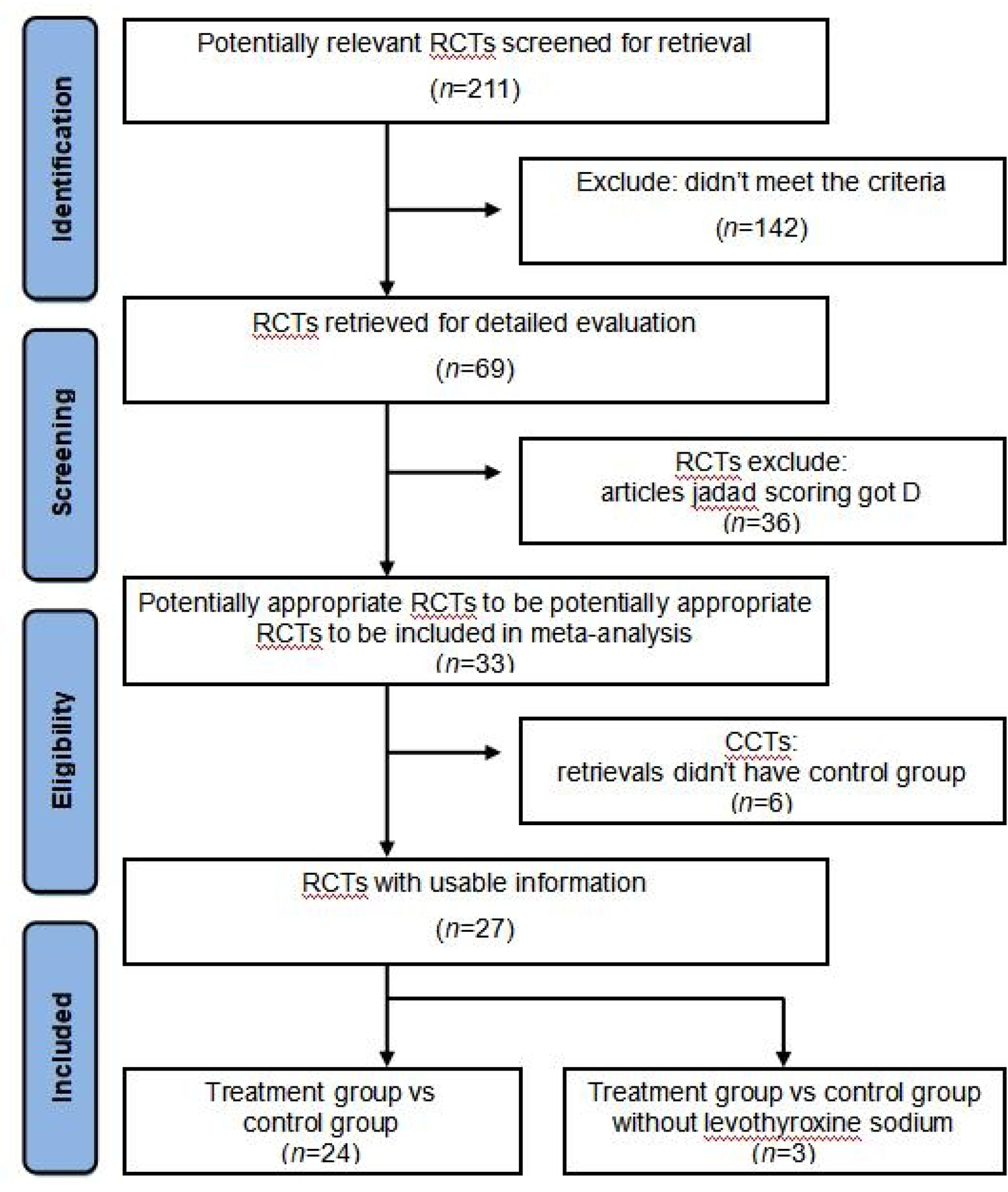
Flow diagram for identification of eligible literatures for this meta-analysis.

**Table 1.**
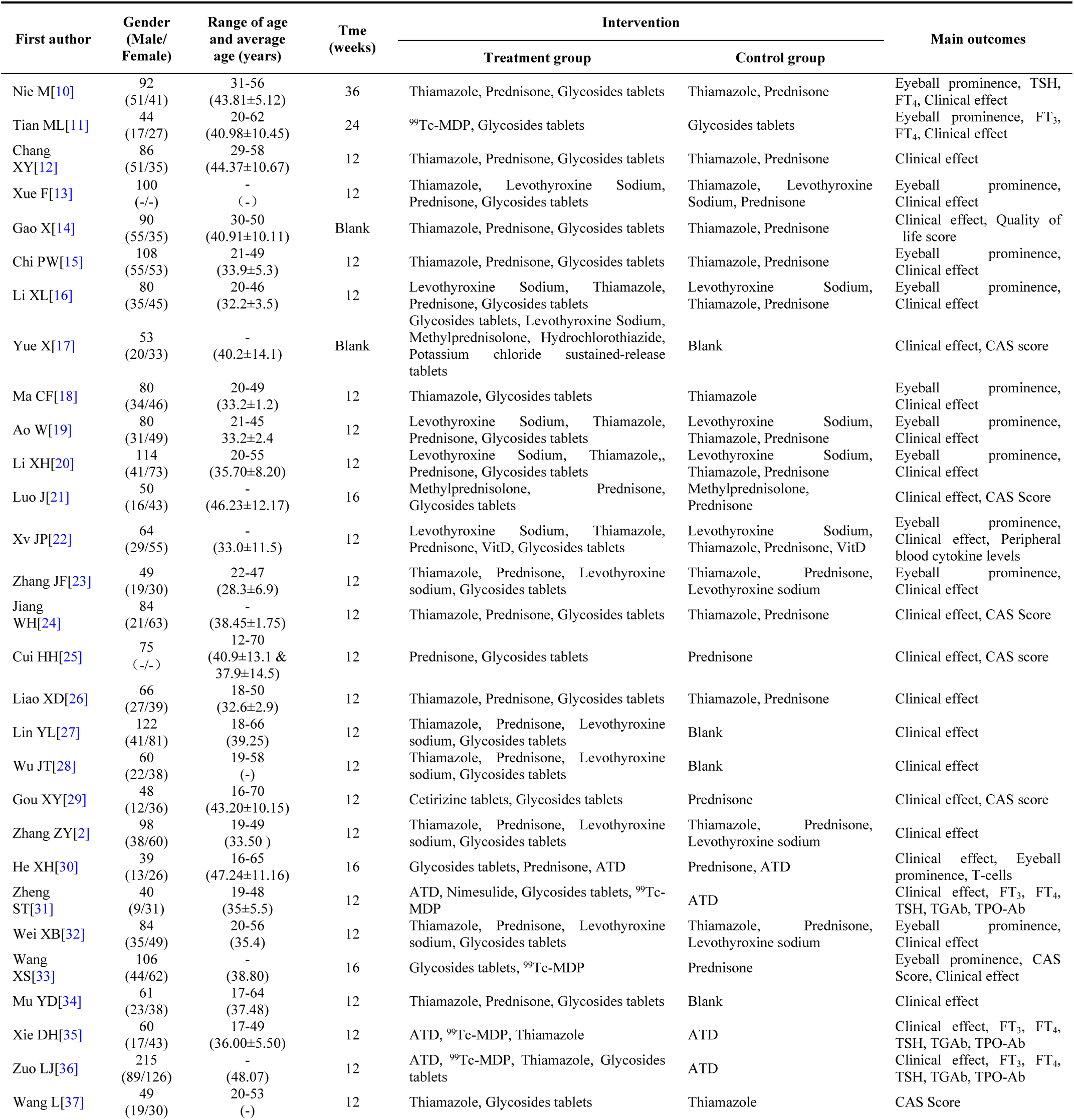
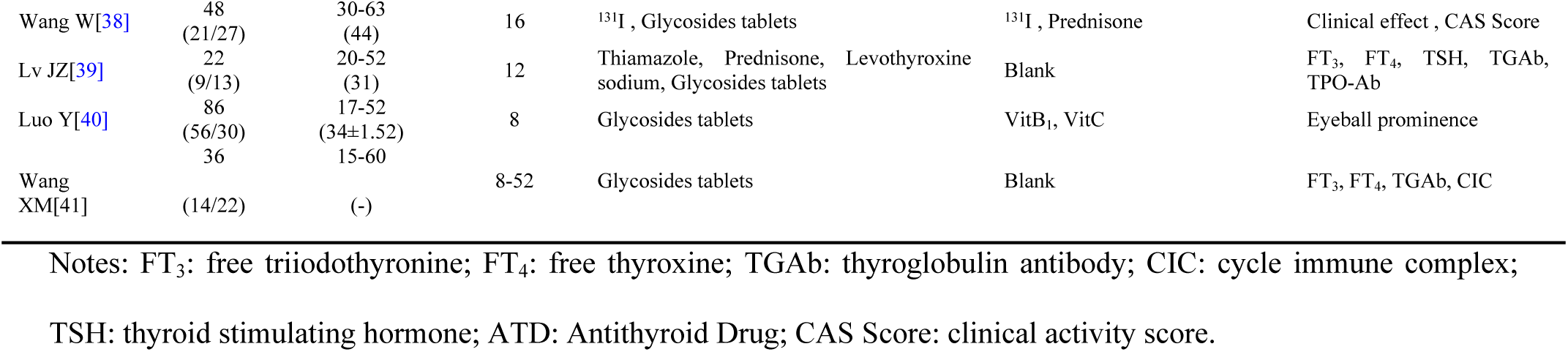
The main characteristics of the trials.

**Table 2.** The methodological quality of the trials.

| First author | Year | City, Province, Country | Random | Blind method | Baseline consistency | Fall off | Follow up | Adverse eventss | Grade |
| --- | --- | --- | --- | --- | --- | --- | --- | --- | --- |
| Nie M[10] | 2021 | Changyuan, Henan, China | Mentioned | Unclear | Consensus | Unclear | Unclear | Unclear | B |
| Tian ML[11] | 2020 | Puyang, Henan, China | Mentioned | Unclear | Consensus | Unclear | Unclear | Unclear | C |
| Chang XY[12] | 2019 | Tieling, Liaoning, China | Mentioned | Unclear | Consensus | Unclear | Unclear | Unclear | C |
| Xue F[13] | 2019 | Weifang, Shandong, China | Mentioned | Unclear | Consensus | Unclear | Unclear | Unclear | C |
| Gao X[14] | 2018 | Lingyuan, Liaoning, China | Unclear | Unclear | Consensus | Unclear | Unclear | Unclear | C |
| Chi PW[15] | 2017 | Xuchang, Henan, China | Mentioned | Unclear | Consensus | Unclear | Unclear | Mentioned | B |
| Li XL[16] | 2017 | Yulin, Shaanxi, China | Mentioned | Unclear | Consensus | Unclear | Unclear | Unclear | C |
| Yue X[17] | 2017 | Zhengzhou, Henan, China | Unclear | None | Consensus | Unclear | Unclear | Mentioned | B |
| Ma CF[18] | 2016 | Bayannur, Inner Mongolia, China | Mentioned | Unclear | Consensus | Unclear | Unclear | Unclear | C |
| Ao W[19] | 2015 | Zhengzhou, Henan, China | Mentioned | Unclear | Consensus | Unclear | Unclear | Unclear | C |
| Li XH[20] | 2015 | Anyang, Henan, China | Mentioned | Unclear | Consensus | Unclear | Unclear | Unclear | C |
| Luo J[21] | 2015 | Enshi, Hubei, China | Mentioned | Unclear | Consensus | Unclear | Unclear | Mentioned | B |
| Xv JP[22] | 2014 | Jinhua, Zhejiang, China | Mentioned | Unclear | Consensus | Unclear | Unclear | Mentioned | B |
| Zhang JF[23] | 2014 | Shaoxing, Zhejiang, China | Mentioned | Unclear | Consensus | Unclear | Unclear | Unclear | B |
| Jiang WH[24] | 2013 | Huangshi, Hubei, China | Mentioned | Unclear | Consensus | Unclear | Unclear | Unclear | C |
| Cui HH[25] | 2013 | Nanjing, Jiangsu, China | Mentioned | Unclear | Consensus | Unclear | Unclear | Mentioned | B |
| Liao XD[26] | 2012 | Leshan, Sichuan, China | Mentioned | Unclear | Consensus | Unclear | Unclear | Unclear | B |
| Lin YL[27] | 2012 | Nanning, Guangxi, China | Unclear | None | Consensus | Unclear | Unclear | Unclear | C |
| Wu JT[28] | 2012 | Jiyuan, Henan, China | Unclear | None | Consensus | Unclear | Unclear | Mentioned | B |
| Gou XY[29] | 2012 | Chongqing, China | Mentioned | Unclear | Consensus | Unclear | Unclear | None | B |
| Zhang ZY[2] | 2010 | Pingdingshan, Henan, China | Mentioned | Unclear | Consensus | Unclear | Unclear | Unclear | B |
| He XH[30] | 2010 | Guiyang, Guizhou, China | Mentioned | Unclear | Consensus | Unclear | Unclear | Mentioned | B |
| Zheng ST[31] | 2010 | Siping, Jilin, China | Mentioned | Unclear | Consensus | Unclear | Unclear | Unclear | C |
| Wei XB[32] | 2009 | Luoyang, Henan, China | Mentioned | Unclear | Consensus | Unclear | Unclear | Unclear | B |
| Wang XS[33] | 2009 | Hengshui, Hebei, China | Mentioned | Unclear | Consensus | Unclear | Unclear | Mentioned | C |
| Mu YD[34] | 2009 | Shangqiu, Henan, China | Unclear | None | Consensus | Unclear | Unclear | Unclear | C |
| Xie DH[35] | 2007 | Zhuhai, Guangdong, China | Mentioned | Unclear | Consensus | Unclear | Unclear | Mentioned | B |
| Zuo LJ[36] | 2007 | Kunming, Yunnan, China | Mentioned | Unclear | Consensus | Unclear | Unclear | None | C |
| Wang L[37] | 2007 | Yurao, Zhejiang, China | Unclear | Unclear | Consensus | 1 case | Unclear | Mentioned | B |
| Wang W[38] | 2004 | Xinxiang, Henan, China | Mentioned | Unclear | Consensus | Unclear | Mentioned | Unclear | B |
| Lv JZ[39] | 2003 | Ningbo, Zhejiang, China | Unclear | None | Consensus | Unclear | Unclear | Mentioned | B |
| Luo Y[40] | 2002 | Taiyuan, Shanxi, China | Unclear | None | Consensus | Unclear | Unclear | Unclear | C |
| Wang XM[41] | 1995 | Yangzhou, Jiangsu, China | Unclear | None | Consensus | Unclear | Unclear | Unclear | C |

### 3.2. Quality assessment

According to the quality evaluation standard for all included RCTs and CCTs for quality assessment and analysis. 11 RCT articles were rated as B grade. 16 articles received a C grade. The evaluation and results are presented in Table 1 and 2. Trials of poor quality (D grade) were excluded.

### 3.3. Results of meta-analysis

33 RCTs or CCTs were published between 2002 and 2021 in China. There were 2,134 cases in 27 RCTs, with 1,104 cases in the treatment group and 1,030 cases in the control group. The treatment group received Tripterygium Glycosides in combination with Thiamazole, Prednisone, Levothyroxine sodium, or a combination of these medications. While the TRIO patients in the control group were treated with Thiamazole, Prednisone, Levothyroxine sodium, or a combination of these treatments. The treatment effect was categorized into four grades: cured, significantly effective, effective, and invalid.

Heterogeneity among studies was assessed using Cochran’s Q test. The *P* value (*P* < 0.01, I^2^ < 0.5) of the Q test < 0.01, a random effect model was used; otherwise, a fixed effect model was used. For each model, the effect summary odds ratio (OR) and its 95% CI were calculated.

Meta-analysis results showed that the overall effectiveness rate of TAO treatment in the treatment group and the control group was *P* = 0.05, I^2^ = 0.33 < 0.5 [MD = 4.45, 95% CI (3.31, 5.99), *P* < 0. 00001], with the former significantly outperforming the latter. At the same time, a risk assessment was conducted for both groups in the study (Fig 2).

**Fig 2.**
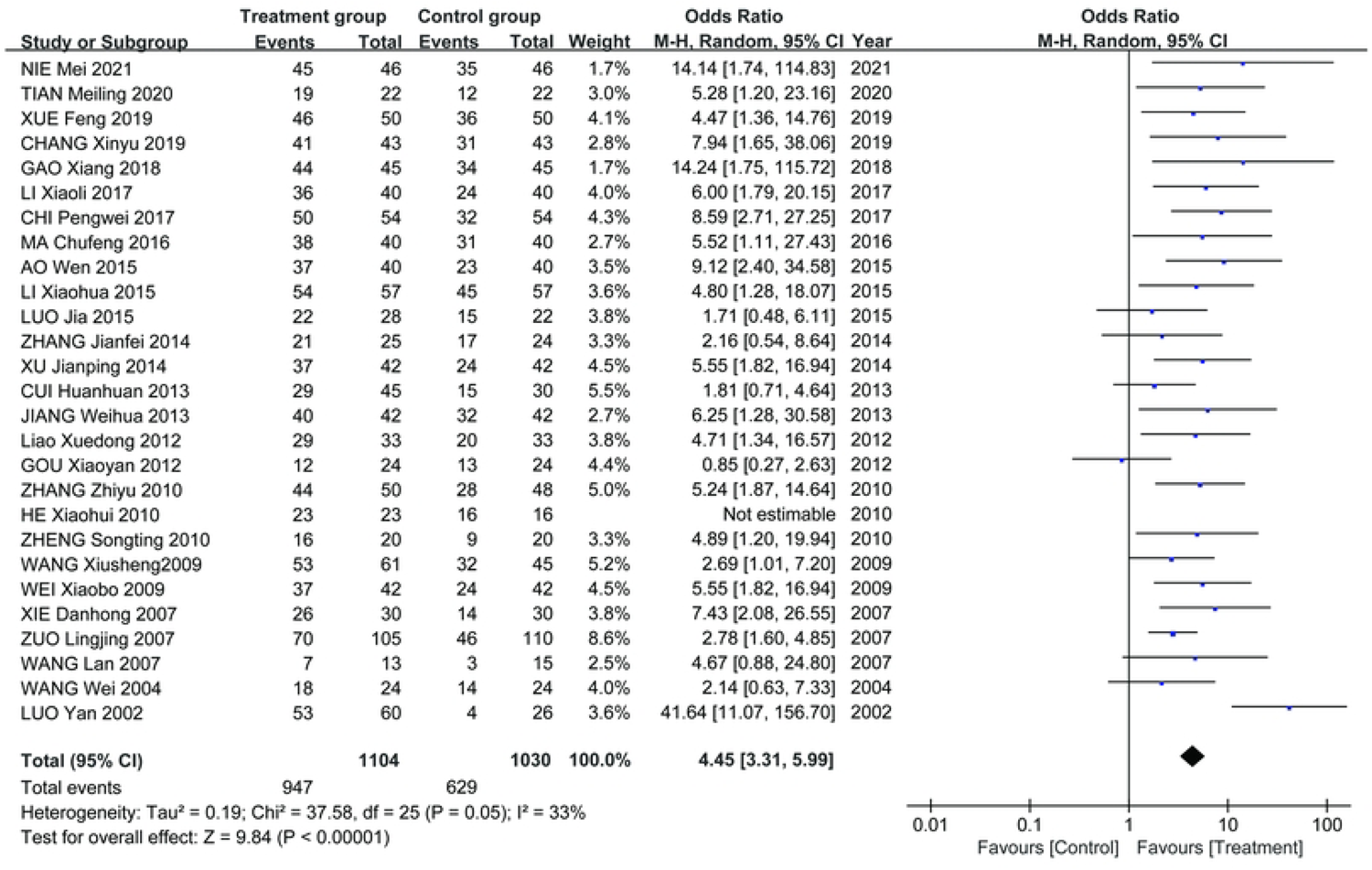
The clinical effectiveness rate.

The clinical effectiveness rate of eyeball prominence was *P* < 0. 00001, I^2^ = 0.98 > 0.5 [MD = 2.40, 95% CI (2.28, 2.51), *P* < 0.00001] (Fig 3).

**Fig 3.**
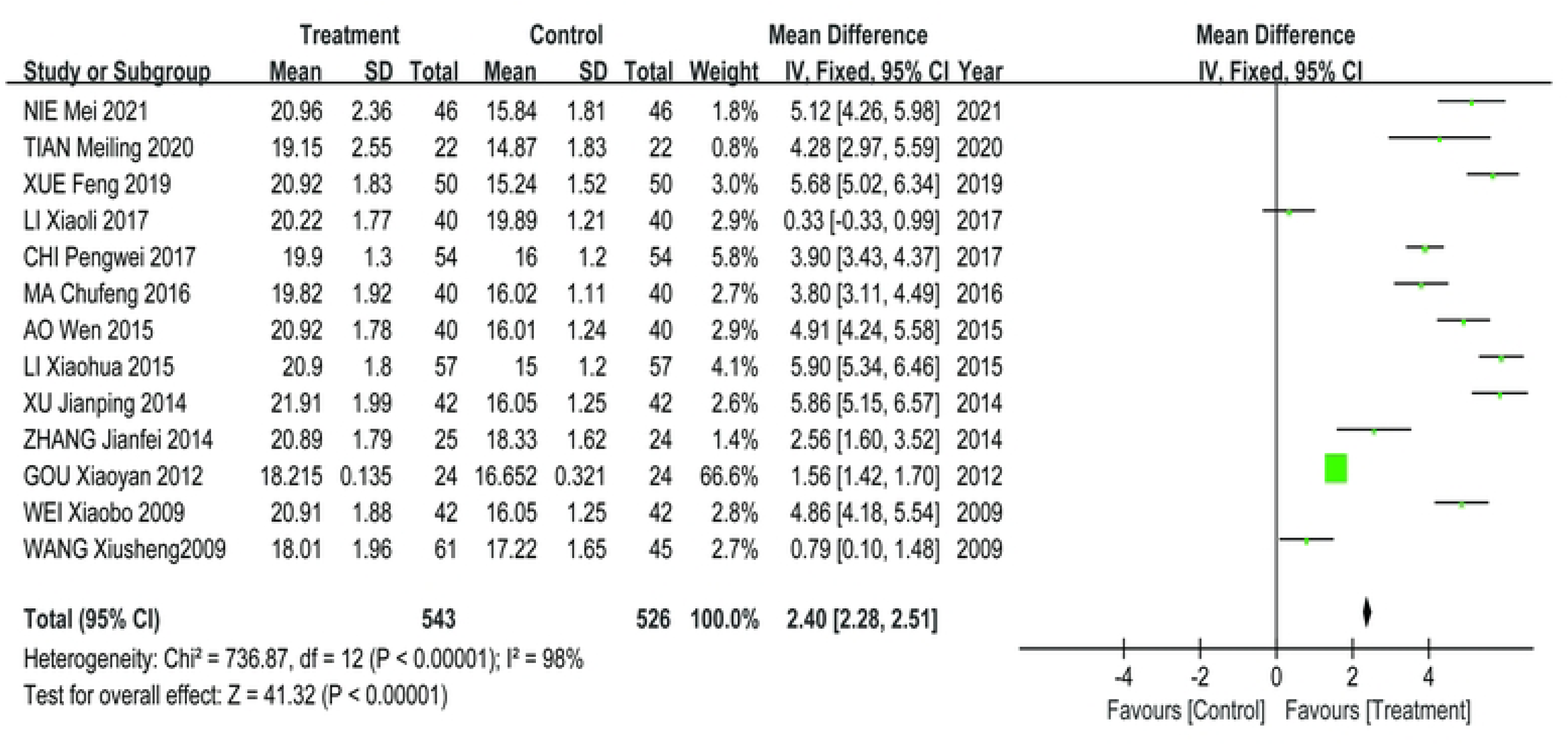
The clinical effectiveness of the eyeball prominence.

The clinical effectiveness rate of CAS score was *P* < 0. 00001, I^2^ = 0.89 > 0.5 [MD = 1.68, 95% CI (1.50, 1.85), *P* < 0.00001] (Fig 4).

**Fig 4.**
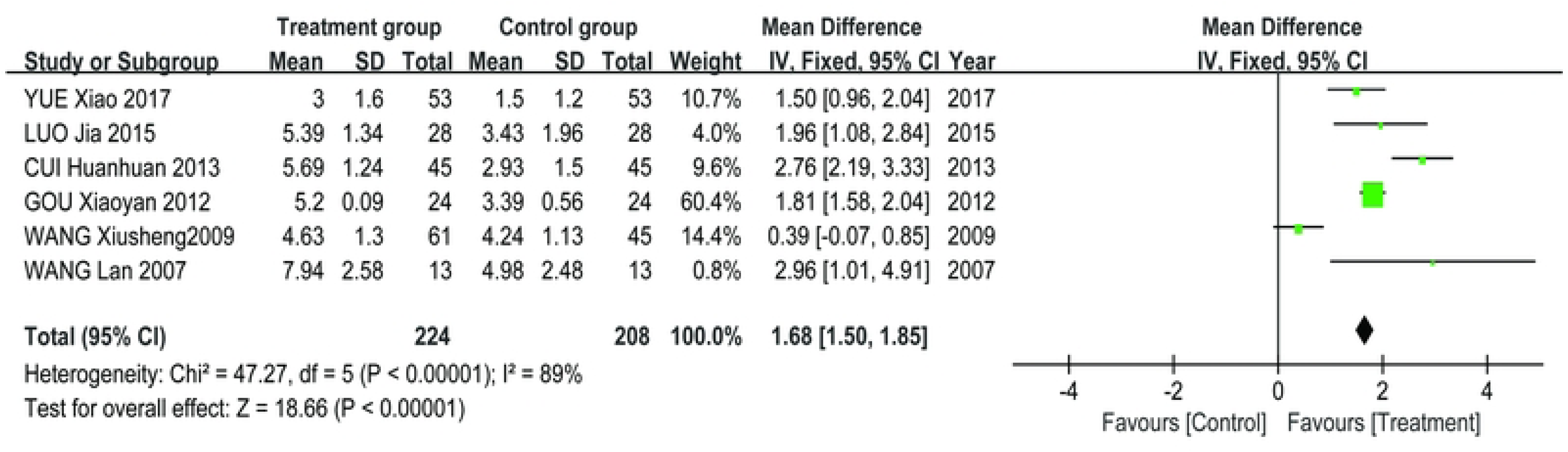
The clinical effectiveness of the eye CAS score.

The clinical effectiveness rate of FT_3_ was *P* < 0. 00001, I^2^ = 0.98 > 0.5 [MD = 0.95, 95% CI (0.81, 1.08), *P* < 0.00001] (Fig 5A), the clinical effectiveness rate of FT_4_ was *P* < 0. 00001, I^2^ = 0.95 > 0.5 [MD = 2.12, 95% CI (1.99, 2.25), *P* < 0. 00001] (Fig 5B), and the clinical effectiveness rate of TSH was *P* < 0.00001, I^2^ = 0.89 > 0.5 [MD = −0.19, 95% CI (−0.21, −0.17), *P* < 0. 00001] (Fig 5C).

**Fig 5.**
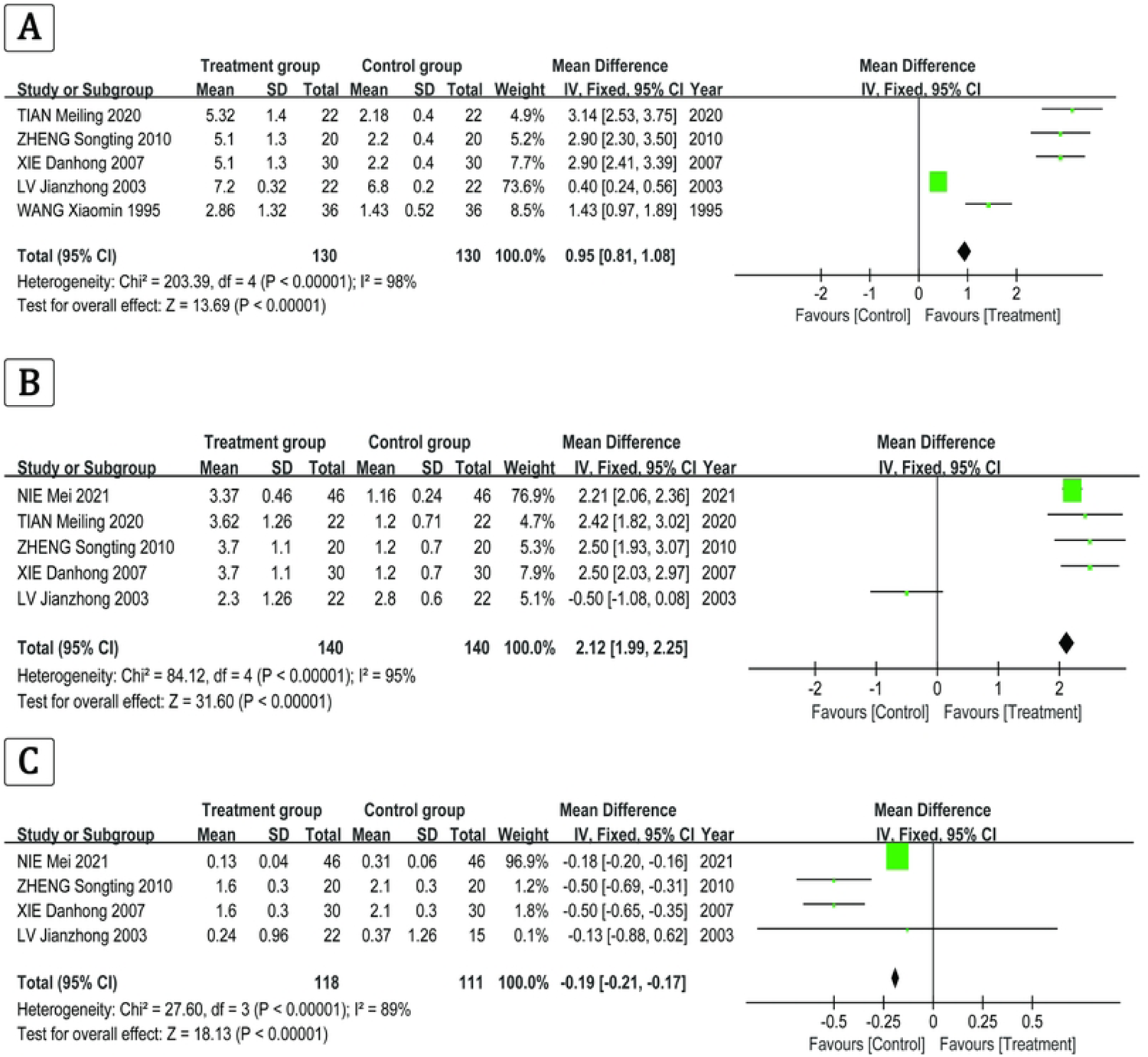
The clinical effectiveness rate of the FT_3_, FT_4_, and TSH.

### 3.4. Adverse events

11 studies reported adverse events. In Chi PW’s study [15], 7 instances of nausea and vomiting, 3 instances of reduced appetite, and 2 instances of diarrhea were reported in the control group. In the observation group, 2 cases of nausea and vomiting, 1 case of reduced appetite, and 1 case of diarrhea were reported. In Yue X’s study [17], adverse reactions mainly occur during hormone shock and the application of somatostatin. Among them, there were 21 cases of abdominal distension, 5 cases of diarrhea, 11 cases of nausea, 5 cases of vomiting, and 3 cases of hypoglycemia, mainly during the use of somatostatin. Additionally, there were 13 cases of abnormal blood glucose: 10 cases of elevated fasting blood glucose levels and 3 cases of early morning hypoglycemia. The elevated fasting blood glucose mainly occurred during the hormone shock treatment period. After 1 week, the fasting blood glucose levels were within the normal range upon re-examination. Exciting insomnia mainly occurs during the hormone pulse therapy period, and the symptoms disappear after the completion of the pulse therapy. There were 4 cases of transient elevated blood pressure, 4 cases of abnormal liver function, and 1 case of hypokalemia. In Luo J’s study [21], 8 cases in the observation group experienced weight gain, elevated blood sugar, elevated blood pressure, and upper abdominal discomfort, respectively. In the control group, 15 cases experienced weight gain, hirsutism, epigastric discomfort, elevated blood glucose, elevated blood pressure, and elevated liver transaminase. In Xv JP’s study [22], the control group had 3 cases of weight gain, 1 case of osteoporosis, and 3 cases of peptic ulcers. In the treatment group, there were 3 cases of mild increases in serum alanine aminotransferase and 3 cases of decreased menstrual flow. In Cui HH’s study [25], 2 cases experienced mild menstrual abnormalities, 2 cases experienced stomach discomfort, 1 case had mild transaminase abnormalities, 1 case gained weight during medication, and exhibited Cushing’s face in the control group. After 1 month of medication, limb muscle stiffness occurred, but no significant changes in blood glucose were observed in all patients. There were 4 cases of mild menstrual abnormalities and 1 case of erythra in treatment group II. In Wu JT’s study [28], 4 cases showed a decrease in WBC, 2 cases experienced gastrointestinal reactions, and 1 case had mild liver dysfunction. All of them recovered after receiving symptomatic treatment. In He XH’s study [30], 3 cases experienced weight gain and hirsutism, while 2 cases experienced acid reflux and upper abdominal discomfort in the treatment group. In Wang XS’s study [33], there were 4 cases of short-term blood glucose elevation and 7 cases of insomnia due to excitement in the control group during the treatment process. The aforementioned side effects gradually disappeared with the decrease in hormone dosage, and no special treatment is needed. In Xie DH’s study [35], a small number of patients experienced vascular pain at the ^99^Tc MDP infusion site. The discomfort symptoms disappeared after the infusion speed was reduced in the treatment group. Some patients experienced weight gain and excessive nighttime urination after receiving low doses of dexamethasone, and no abnormalities were found in routine blood and urine tests. After the treatment, and routine blood and urine tests did not reveal any abnormalities. In Wang L’s study [37], 1 woman experienced amenorrhea and withdrew from the observation. However, she recovered after discontinuing the medication. In Lv JZ’s study [39], 21 cases experienced anorexia, nausea, vomiting, and diarrhea, and their symptoms were relieved.

### 3.5. Risk of bias within studies

Publication bias analysis was conducted on these 27 pieces of literature using a funnel plot, which resulted in a symmetric distribution. Begg’s test and Egger’s test were also conducted. Both of the *P* values were > 0.05, indicating that there was no publication bias in the included trials. All the matching points were found within the 95% CI.

The bias was evaluated by conducting a funnel plot analysis of the Tripterygium Glycosides treatment for TAO. The accuracy improved as the sample size increased. The amount of literature included is insufficient, scattered within the pyramid, and symmetrically distributed alongside the axis, indicating minimal bias (Fig 6A). The points corresponding to the CAS score (Fig 6C), FT_4_ (Fig 6E), and TSH (Fig 6F) in the included trials are primarily situated within the 95% CI, with a scattered distribution within the range, basically symmetrical on both sides, and presenting a funnel-shaped shape, indicating a small publication bias in the trials included. The eyeball prominence (Fig 6B) corresponding points were outside the range of the 95% CI. This may be attributed to factors such as small sample studies, non-significant results not being published or cited. The corresponding point of FT_3_ (Fig 6D) is asymmetric on both sides of the axis.

**Fig 6.**
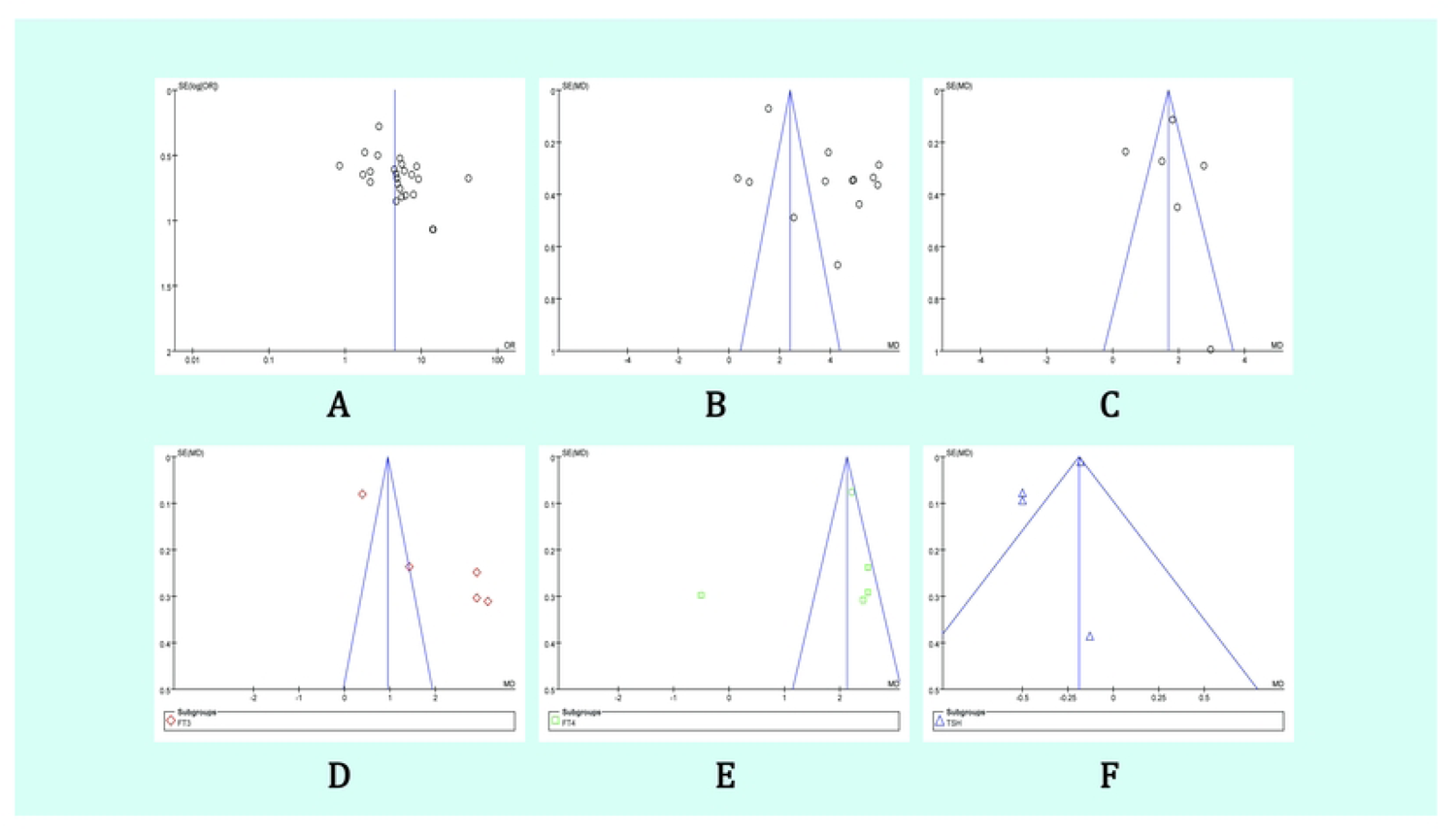
Funnel plot of the literatures analyzed in the meta–analysis.

## 4. Discussion

TAO is a multifactorial ocular disorder caused by thyroid disease, which often manifests as eye redness, eye pain, exophthalmos, edema, and impaired movement of periocular muscles, etc. 40%-70% of TAO patients suffer from hyperthyroidism. Currently, TAO is clinically categorized into two types: one is ocular infiltrative type, also known as endocrine ophthalmoplegia or malignant Graves’ ophthalmopathy, which accounts for 5%-10% of patients with hyperthyroidism. The other is the non-infiltrative type of ophthalmoplegia, also known as simple or benign exophthalmoplegia, which is usually caused by sympathetic stimulation of the periorbital or upper facial muscles. Ocular infiltrative TAO is an autoimmune disease caused by hyperplasia, lymphocytic infiltration and edema of the retro-ocular tissues and is influenced by a variety of factors, such as smoking, genetics and the environment [18,44].

The early pathologic changes in these diseases are the infiltration of lymphocytes and serum cells into the periocular muscles and connective tissues [45–47].

During the course of the disease, there is a buildup of collagen in the periocular muscles, which leads to fibroblasts and fat cell deposits, the presence of which is of greater significance because it indirectly confirms that the disease undergoes a longer and slower progression. The presence of fat deposits is more significant because it indirectly confirms that the disease undergoes a longer and slower progression. Current research suggests that the correlation between thyroid disease and ocular symptoms may explain the following findings: (1) edema leading to an increase in the volume of the contents of the eye sockets; (2) the production of hydrophilic glucosamines and peptidoglycans; and (3) an accumulation of adipose tissue in the eye sockets.

At the present time, the combined use of tretinoin preparations for the treatment of infiltrative TAO is widely used in the clinic. This study found that treatment with Tripterygium Glycosides may be more effective in enhancing the response rate, quality of life, and FT_3_ levels compared to treatment with Prednisone, Levothyroxine sodium, and/or Thiamazole alone. Although adverse reactions were still present in the control group, overall the rate of adverse events was lower in the observation group and the clinical benefit was much higher than in the control group. At the same time, a large body of medical evidence shows that Tripterygium Glycosides is effective in the treatment of eye protrusion in hyperthyroidism. It can inhibit cellular and humoral immunity and improve the immune status of the body. It has also been shown to inhibit the formation of self-antigens in the tissues behind the eye, thereby reducing eye protrusion [48].

Tripterygium Glycosides is a traditional herbal medicine that originated from China. It has been widely used in China for the treatment of various diseases, including rheumatic diseases, skin diseases, and diabetic nephropathy. In recent years, there have been international studies conducted on Tripterygium Glycosides by renowned institutions such as Harvard University and the University of California, Los Angeles (U.S.A.), the Institute of Pharmacology and Osaka University (Japan), the Siberian Branch of the Russian Academy of Sciences and the Russian Federal Institute of Medical and Biotechnological Research (Russia), as well as the Indian Academy of Medical Sciences and the University of Delhi (India). The research conducted in these countries has mainly focused on studying the pharmacological effects, clinical applications, quality control, and other aspects of Tripterygium Glycosides.

Through systematic analysis, we found that the clinical results of applying Tripterygium Glycosides were significantly better than those of the control group. This fully explains the safety, reliability, and precise clinical efficacy of the use of Tripterygium Glycosides in the treatment of hyperthyroid eye protrusion. In addition, Tripterygium Glycosides may improve the efficacy of the basic treatment and may lead to a reduction of the drug dose or complete discontinuation of the treatment. Based on these characteristics, Tripterygium Glycosides treatment may be an ideal solution for hyperthyroidism-like herniated eye disease.

It is important to note that long-term use of Tripterygium Glycosides can cause some damage to various body systems. For example, long-term use of Tripterygium Glycosides at higher-than-average doses can lead to reversible liver and kidney damage, while approximately 20% of patients experience gastrointestinal reactions such as loss of appetite, nausea, vomiting, abdominal pain, diarrhea, or constipation. As for the hematopoietic system, the effects of Tripterygium Glycosides use are mainly manifested in the form of a decrease in the number of white blood cells and platelets included [49,50]. It has even been found that long-term use of Tripterygium Glycosides causes skin and mucosal reactions such as oral mucosal ulcers, dryness of the mouth and eyes, roughness and dryness of the skin, rashes, skin sclerosis, and increased melanin production, which is usually associated with the inhibition of the IL-23/IL-17 pathway [51,52]. In addition, long-term use of Tripterygium Glycosides can inhibit ovarian function and cause menstrual disorders such as decreased menstrual flow or amenorrhea in women, and in men, it may lead to a decrease in sperm count or sperm motility [53,54].

Despite the inevitable problems associated with the use of Tripterygium Glycosides in the treatment of disease, its benefits in the treatment of various diseases cannot be ignored. Currently, there are several meta-analyses of interest due to the efficacy of Tripterygium Glycosides in the treatment of renal diseases, dermatological disorders, rheumatoid arthritis, and nephrotic syndrome ^55-58^. Therefore, it is crucial to conduct a multicenter randomized, double-blind clinical trial to study the efficacy of trehalose in TAO [59,60].

## 5. Conclusion

This meta-analysis demonstrates that the experience with the treatment of TAO using Tripterygium Glycosides was promising. The existing evidence suggests that treatment with Tripterygium Glycosides may be more effective in enhancing the response rate, quality of life, and FT_3_ levels compared to treatment with Prednisone, Levothyroxine sodium, and/or Thiamazole alone.

## Data Availability

All relevant data are within the manuscript and its Supporting Information files.

## Author contributions

Conceptualization: Mingzhe Li, Tianshu Gao.

Data curation: Mingzhe Li, Bingchen Wei.

Formal analysis: Bingchen Wei, Chenghan Gao.

Funding acquisition: Mingzhe Li, Tianshu Gao.

Investigation: Mingzhe Li, Bingchen Wei, Chenghan Gao.

Methodology: Mingzhe Li, Tianshu Gao.

Writing-original draft: Mingzhe Li, Bingchen Wei.

Writing-review: Mingzhe Li, Bingchen Wei, Chenghan Gao, Tianshu Gao.

## References

1. Ying JM. Progress of Chinese medicine in treating hyperthyroid exophthalmos. Journal of Traditional Chinese Medicine.25:979–981(2009).

2. Zhang ZY, Wang JH. Tripterygium Glycosides with tapazole and prednisone in the treatment of hyperthyroidism exophthalmos. Journal of Medical Forum. 31 (8): 110–111(2010).

3. Webb MS, John stone S, Morris TJ, Kennedy A, Gallagher R, Harasym N, et al. In vitro and in vivo characterization of a combination chemotherapy formulation consisting of vinorelbine and phosphatidyl bine. European Journal of Pharmaceutics and Bio pharmaceutics. 65 (165): 289–299(2010).

4. Li XW. Clinical observation of 50 cases of children with purpura nephritis treated with Tripterygium Glycosides tablets. Jiangsu Medical Journal. 12: 664–665(1987).

5. Ministry of health of the people’s Liberation Army General Logistics Department. Diagnosis of clinical diseases based on the improvement of the standard. Beijing: People’s military medical press. 1198–1991(1987).

6. Ministry of Health, PRC. Guiding principle of clinical research on new drugs of Traditional Chinese Medicine. Beijing: The Medicine Science and Technology Press of China. section 1: 168(1993).

7. Wiersinga WM, Prummel MF. Grave’s ophthalmopathy: a rational approach to treatment. Trends Endocrinal Metabolism. 13 (7): 280(2002).

8. Gu MJ, Wu WY, Liu CH, Liu XL, Li X, Liu ZM. Efficacy of prednisone in the treatment of thyroid associated ophthalmopathy on adrenocortical function. Journal of Xi’an Jiaotong University: Medical Sciences. 24 (2): 177–178(2003).

9. Higgins JPT, Green S. Cochrane Handbook for Systematic Reviews of Interventions Version 5.3 [updated March, 2015].The Cochrane Collaboration,(2015).

10. Nie M. Efficacy evaluation of Tripterygium wilfordii polyglycoside combined with Thiamazole and prednisone acetate tablets in the treatment of hyperthyroid exophthalmos. Heilongjiang Journal of Traditional Chinese Medicine. 50(3):162–163(2021).

11 Tian ML. Therapeutic effect of Tripterygium wilfordii polyglycoside combined with technetium 99-methylenediphosphonate on thyroid associated ophthalmopathy. Practical Clinical Journal of Integrated Traditional Chinese and Western Medicine. 20(5):17–18(2020).

12. Chang XY. Effect of Tripterygium wilfordii polyglycoside combined with tapazole and prednisone on hyperthyroidism exophthalmos. Guide of China Medicine, 17(21):167–168. DOI:10.15912/j.cnki.gocm.21.129.(2019).

13. Xue F, Zhang WJ. Analysis of the symptom and outcome of hyperthyroidism exophthalmos treated with Tripterygium wilfordii glycosides. World Latest Medicine Information. 19(46):214. DOI:10.19613/j.cnki.1671-3141.2019.46.127(2019).

14. Gao X. Evaluation of the clinical efficacy of Tripterygium wilfordii polyglycoside, tapazole and prednisone in the treatment of hyperthyroid exophthalmos. Guide of China Medicine. 16(2):187–188. DOI:10.15912/j.cnki.gocm.2018.02.157(2018).

15. Chi PW. Clinical analysis of Tripterygium wilfordii polyglycoside combined with Thiamazole and prednisone in the treatment of hyperthyroid exophthalmos. Journal of North Pharmacy. 14(9):34–35(2017).

16. Li XL, Ma T. Clinical effect of tripterygium glycosides combined with tazobactam and prednisone on patients with hyperthyroid exophthalmos. Clinical Research and Practice. 2(22):75–76. DOI:10.19347/j.cnki.2096-1413.201722037(2017).

17. Yue X, Wang YY, Yang Y, Wen SM, Sun LG. Clinical efficacy and safety analysis of glucocorticoids combined with somatostatin and Tripterygium wilfordii glycosides in the treatment of thyroid associated ophthalmopathy. Henan Medical Research. 26(1): 31–33(2017).

18. Ma CF, Wang YS. Clinical efficacy of Tripterygium wilfordii glycosides combined with Methimazole in the treatment of hyperthyroid exophthalmos. World Latest Medicine Information. 16(75):104(2016).

19. Ao W. Observation on the therapeutic effect of combination therapy of Tabazole, prednisone, and Tripterygium wilfordii glycosides in patients with hyperthyroidism exophthalmos. Medical Journal of Chinese People’s Health. 27(13):95–96(2015).

20. Li XH. Clinical analysis of Tripterygium wilfordii glycosides combined with Tabazolidinolone in the treatment of hyperthyroidism exophthalmos. Clinical Research. 23(5): 60–61(2015).

21. Luo J, Huang J, Ye M. The observation of curative effect of glucocorticoids combined with Glucosida Tripterygii TOTA in the treatment of Graves’ ophthalmopathy. Practical Journal of Clinical Medicine. 12(5): 174–176(2015).

22. Xv JP, Xv C, Chen J, Jin ZH, Zheng HF, Zhu J. Graves The level of cytokines in peripheral blood of ophthalmopathy and the effect of Tripterygium wilfordii polyglycoside intervention. China Journal of Chinese Materia Medica. 39(3): 544–547(2014).

23. Zhang JF, Kong YZ, Pan HZ. Clinical efficacy of the treatment of hyperthyroidism exophthalmos with Tripterygium Glycosides tablets. Chinese Rural Health Service Administration. 6: 761–762(2014).

24. Jiang WH. Clinical observation of Tripterygium wilfordii polyglycoside combined with tapazole and prednisone in the treatment of hyperthyroidism exophthalmos. World Health Digest. 10(22): 178–179(2013).

25. Cui HH, Ye XZ, Li YL, Lu B, Peng L, Xv YX, et al. Clinical efficacy of mycophenolate mofetil and other Immunosuppressive drug in the treatment of thyroid associated ophthalmopathy. Chinese Journal of Clinicians (Electronic Edition). 7(24):11197–11200(2013).

26. Liao XD. Clinical observation of hyperthyroidism exophthalmos treated with Tripterygium Glycosides combined with methimazole and prednisone. China Health Care and Nutrition. 10: 1514–1515(2012).

27. Lin YL. Treatment 122 cases of hyperthyroidism exophthalmos treated with Tripterygium Glycosides tablets combined with methimazole and prednisone. China Journal of Pharmaceutical Economics. 7 (5): 119–120(2012).

28. Wu JT. Clinical analysis of hyperthyroidism exophthalmos treated with Tripterygium Glycosides tablets combined with methimazole and prednisone. Contemporary Medicine. 18 (22): 79–80(2012).

29. Gou XY, Cheng G. Clinical Study on Certirizine Combined Tripterygium Glycosides for Treating 24 Cases of Thyroid Associated Ophthalmopathy. China Pharmaceuticals. 21(18):85–86(2012).

30. He XH, Kong DM. Tripterygium wilfordii polyglycosides combined with low-dose prednisone in the treatment of 23 cases of Graves’ ophthalmopathy. New Chinese Medicine. 42(8): 65–66(2010).

31. Zheng ST, Zhang B, Mei F. Multiple enhanced immunosuppression for hyperthyroid exophthalmos: Yunke, Nimesulide, Tripterygium wilfordii polyglycoside tablets for Graver ophthalmopathy. World Health Digest. 7(23): 65–66(2010).

32. Wei XB. The effect of Tripterygium Glycosides tablets on the treatment of hyperthyroidism exophthalmos. China Modern Doctor. 20: 97+101(2009).

33. Wang XS, Li GL, Wang Q, Ge CJ, Zhang L. Clinical study of Yunke combined with Tripterygium wilfordii glycosides in the treatment of thyroid associated ophthalmopathy. Chinese Journal of Practical Ophthalmology. 27(12):1369–1371(2009).

34. Mu YD. Case analysis of Tripterygium wilfordii united tapazole prednisone in the treatment of 61 cases of thyrotoxic exophthalmoses. China Medical Herald. 6 (23): 25–27(2009).

35. Xie DH, Sun L, Shu XC, Ye LH, Shen J, Lu HY. Combination of technetium [^99^Tc] methylenediphosphonate, dexamasone and tripterygium glucosides in treatment of Graves ophthalmopathy. Chinese Journal of New Drugs and Clinical Remedies. 26(12): 905–908(2007).

36. Zuo LJ, Yang JS. Observation on the therapeutic effect of ^131^I combined with Tripterygium wilfordii glycosides on Graves’ ophthalmopathy. China Health Care. 15(19): 51–52(2007).

37. Wang L. Immunosuppressive drug therapy for Graves ophthalmopathy. Zhejiang Clinical Medicine Journal. 9:197(2007).

38. Wang W, Yang B, Sun HJ, Zhou YL. Clinical study of ^131^I and glucoside tripterygium total tablets on Graves’ophthalmopathy. Chinese Journal of Nuclear Medicine. 24(03): 38–39(2004).

39. Lv JZ. Prednisone and Tripterygium Glycosides tablets in the treatment of hyperthyroidism exophthalmos. Zhejiang Journal of Integrated Traditional Chinese and Western Medicine. 13 (5): 299–230(2003).

40. Luo Y, Zheng DW, Wang X, He YQ, Li DP. Clinical Observation of Tripterygium wilfordii Polysaccharide Tablets in the Treatment of Thyroid related Orbital Lesions. China Journal of Chinese Ophthalmology. 12(2):95–97(2002).

41. Wang XM.Clinical study on the treatment of hyperthyroidism exophthalmos with Glycosides tablets. Journal of Jiangsu TCM. 16 (10): 41–42(1995).

42. Shen ZY. The summary and analysis of the influence of Traditional Chinese Medicine on immune function. Chinese Journal of Integrated Traditional and Western Medicine. 12:443–6(1992).

43. Bartalena L, Piantanida E, Gallo D, Lai A, Tanda ML. Epidemiology, Natural History, Risk Factors, and Prevention of Graves’ Orbitopathy. Front Endocrinol (Lausanne).11:615993. doi: 10.3389/fendo.2020.615993(2020).

44. Bai Y. Thyroid disease-basic and clinical. Beijing: Science and Technology Literature Press. 466–476(2002).

45. Shi FX. Observation on the effect of treating Graves eye disease with Tripterygium Glycosides tablets. New Chinese Medicine. 21:472–473(1990).

46. Yang JH, Duan JG. The progress of hyperthyroidism exophthalmos with the treatment of Chinese medicine treatment. New Journal of Traditional Chinese Medicine. 35:74(2003).

47. Li GM, Li GM, Chen LD. A comparative observation on hyperthyroidism exophthalmos with clinical combination of Traditional Chinese and Western Medicine. Chinese Journal of Information on Traditional Chinese Medicine. 15:62(2008).

48. Zheng JR, Lv Y. Clinical and experimental studies of Tripterygium. Journal of Traditional Chinese Medicine. 23:74–78(1982).

49. Liu F. Pharmacological research and clinical application of Tripterygium Glycosides tablets. Chinese Traditional Patent Medicine. 24:385(2002).

50. Li LX, Jin RM, Li Yikui, Fu SG, Huang J, Zhu ZL, et al. Study on the immunosuppressive effect and safety range of multiple dosing of Tripterygium Glycosides tablets. Chinese Journal of New Drugs and Clinical Remedies. 25:248(2006).

51. Chu WL, Kang ZF, Chan SL, Li WY, Liu J. Research progress on inhibition of ocular neovascularization by Chinese herbal monomers. Journal of Traditional Chinese Ophthalmology. 29:166–169(2019).

52. Qin TY, Gao SS, Wang WZ. The inhibitory effect of Tripterygium wilfordii red pigment on the secretion of IL-17 by peripheral blood mononuclear cells in patients with sympathetic ophthalmitis. Chinese Journal of Ocular Fundus Diseases. 34:51–54(2018).

53 Li M, Li Y, Xiang L. Efficacy and safety of Tripterygium glycosides as an add-on treatment in adults with chronic urticaria: a systematic review and meta-analysis. Pharm Biol. 61:324–336. doi: 10.1080/13880209.2023.2169468(2023).

54 Xie D, Li K, Ma T, Jiang H, Wang F, Huang M, et al. Therapeutic Effect and Safety of Tripterygium Glycosides Combined With Western Medicine on Type 2 Diabetic Kidney Disease: A Meta-Analysis. Clin Ther. 44:246–256.e10. doi: 10.1016/j.clinthera(2022).

55. Li WW, Liu XL, Wu H, et al. Efficacy and Safety of Tripterygium wilfordii Hook.F for IgA Nephropathy: A Meta-analysis. Chin J Evid-based Med. 15:206–214(2015).

56. Zhang XM, Xiang SM. Analysis of randomized clinical trials of oral GTT tablets in the treatment of psoriasis vulgaris. Guide of China Medicine. 27:392–393(2013).

57. Wang JQ, Li GC, Zhou XP, Ma Z. Tripterygium wilfordii extraction for treating rheumatoid arthritis: a meta-analysis. Modern Journal of Integrated Traditional Chinese and Western Medicine. 23:1032–1036(2014).

58. Huang Q, Zeng QM, Zheng YH, Xiong JL. Meta Analysis of Tripterygium wilfordii Combined Glucocorticoid therapy in NS Patients. Journal of Liaoning University of TCM. 17:145–149(2015).

59. Sun ZQ. Medical statistics. Beijing: People’s Health Publishing House.623(2002).

60. Wang JY. Evidence based medicine and clinical practice. Beijing: Science press.118–121(2002).

